# Assessing the effectiveness of a pediatrician-led newborn parenting class on maternal newborn-care knowledge, confidence and anxiety: A nonrandomized controlled trial

**DOI:** 10.1101/2019.12.18.19014936

**Authors:** Aileen Gozali, Sherika Gibson, Lianna R. Lipton, Aliza W. Pressman, Blair S. Hammond, Dani Dumitriu

## Abstract

The postpartum hospital stay is a unique opportunity for clinicians to educate parents on the importance of promoting early child development. Pediatricians are well positioned to address both medical and developmental concerns during critical periods of development, yet very few parenting interventions are led by pediatricians.

**Aims:** To assess the impact of a novel one-hour long pediatrician-led Newborn Class on maternal knowledge, confidence, and anxiety.

**Methods:** We conducted a nonrandomized controlled trial to evaluate the effectiveness of the Newborn Class. First-time mothers who delivered a full-term singleton vaginally with no major complications and attended the class were recruited. Mothers who expressed a desire to attend the class but were discharged before a class was offered served as controls.

**Outcome measures:** Maternal self-perceived anxiety and confidence were measured using standardized scales (State-Trait Anxiety Inventory for Adults and Karitane Parenting Confidence Scale). Knowledge on new-born care was assessed using a novel scale.

**Results:** A total of 84 participants (intervention n=36, control n=48) were included in the study. Mothers who attended class showed significantly higher levels of knowledge compared to the control group as well as significantly higher parenting confidence levels. No change was observed in the overall level of anxiety.

**Conclusions:** A short pediatrician-led parenting intervention can be an effective tool in improving maternal confidence and newborn care knowledge. Given the importance of the newborn period in establishing healthy developmental trajectories, this cost- and time-effective intervention could be widely implemented to promote early strong mother-infant relationships that foster healthy development.

## Introduction

Experiences during sensitive periods of development provide scaffolding for a child’s physical, cognitive and behavioral health trajectories (1). During the first years of life, the brain undergoes rapid development and is most vulnerable to external experiences (2). Responsive relationships with care-givers are essential for optimal development, as reflected in better executive functioning (3), better attention (both short term and later in life) (4)(5), fewer addictive behaviors (6), and lower risks of cardiovascular disease in adulthood (7). The verbal stimulation associated with responsive caregiving is positively associated with children’s language skills (8)(9). Responsive parenting can also buffer against the detrimental effects of early childhood adversity, which include poorer cognitive and socio-emotional development (10)(11) and worse health and behavioral outcomes in adulthood, such as obesity and drug use (12)(13).

Specific behaviors of caregivers have been identified as responsive parenting. These include consistent responsiveness, sensitivity to distress, verbal scaffolding, and autonomy support (14)(15)(16)(17). Numerous parenting interventions promoting responsive parenting behaviors have been designed, implemented and evaluated. Meta-analysis of existing interventions shows large variations in methods of delivery (group versus individual), class setting (primary care setting versus community setting), intensity (frequency and duration), cost, and class instructors (18)(19). Although comparisons of class effectiveness have not been made for different methods of delivery, longer interventions were associated with increased parental confidence. Financial and staffing challenges have led to the increased use of pediatric primary care setting as the site for intervention due to its established infrastructure and the already well-attended well-child visits (20)(21); however, very few of them are taught by pediatricians. Yet, pediatricians are uniquely suited to address both medical and developmental topics. Not only do pediatricians have access to children and their families at critical developmental timepoints, but research has shown that parental behaviors, such as daily reading and breast feeding, are more likely to be carried out by parents if they are encouraged by a pediatrician (22). Pediatricians are thus ideally positioned to positively affect the mother-infant relationship (23).

Within this context, the Mount Sinai Parenting Center developed the Newborn Parent Education and Discharge Class (hereafter referred to as the Newborn Class). The class is one-hour long, taught by pediatric attendings and residents, and offered at no-cost to families in the Well Baby Nursery (WBN) at Mount Sinai Hospital. With 98.39% of babies in the U.S. born in hospitals (24), postpartum stay in the hospital is a unique opportunity for clinicians to educate parents on the importance of promoting early child development. Parents are encouraged to attend the Newborn Class during the mother’s postpartum hospital stay. The class covers both medical topics, such as safe sleep, feeding and bathing, as well as developmental topics, such as language development, responsive parenting, and soothing a baby. The class is in addition to the individualized discharge instructions provided by the clinical staff. The Newborn Class aims to promote strong mother-child relationships and increase the skills and support of mothers, with the goal of positively impacting a child’s developmental trajectory.

Currently, the class is offered two to three times per week based on available resources. Here, we present initial data on the effectiveness of the Newborn Class on several maternal outcomes at time of discharge. To evaluate the effectiveness of the Newborn Class, a non-randomized controlled trial was implemented. Mothers who attended the Newborn Class were asked to participate in a questionnaire-based survey at the time of discharge, one day after attending the class. Because the class is available only on certain days of the week, it is inaccessible to a subpopulation of mothers who would attend if it were available. These mothers were case-matched to serve as the controls for the intervention group. Post-intervention knowledge was quantified using a scale developed to specifically target components taught in the Newborn Class. In addition to knowledge, parenting behaviors are strongly affected by parental anxiety and self-efficacy (25)(26)(27). These are commonly assessed measures of parenting intervention for which many validated and standardized scales exist (28). Scales included in the study were chosen to minimize participants’ time commitment.

## Methods

### 2.1. Ethics Statement

The study was conducted in accordance with Mount Sinai’s Institutional Review Board (IRB). Because no personal identifiers were collected during the trial, it qualified for IRB exempt status. Trial registration: ClinicalTrials.gov Identifier: NCT04121390.

### 2.2. Study Design

The study was conducted at the Klingenstein Pavilion of Mount Sinai Hospital located in New York, NY. Mothers who desired to attend the New-born Class were self-selected into control and intervention group based on whether a class was available during their postpartum stay. For the intervention group, a research assistant collected the room numbers of mothers who attended the class. Another research assistant screened for eligible participants and recruited them into the study the next morning, prior to discharge. During the remaining days of the week, mothers were screened and recruited into the control group on the morning of their discharge. Immediately after recruitment and consent, participants in both groups took part in a 10-minute survey measuring the primary outcomes of the study which include anxiety level, confidence level and newborn care knowledge. The study consent form and surveys are provided in **Appendix A**.

### 2.3. Subjects

Mothers were recruited between May, 2018, and August, 2019. Inclusion criteria included: 1) first-time mother, 2) above 18 years old, 3) vaginal delivery of a healthy full-term singleton, 4) no major perinatal complications (per self-report), 5) English-speaking, and 6) consent to participate. These inclusion criteria were designed to control for known factors associated with parental anxiety and confidence (29)(30).

### 2.4. The intervention

The Newborn Class is taught in the lounge of the Well Baby Nursery (WBN). On days the class is taught, parents receive reminders through overhead announcements and from nurses and residents. The class is open to all parents. All parents in the WBN, regardless of whether or not they attend the class, are given links to online videos with demonstrations by pediatricians at Mount Sinai covering similar topics to what is taught in the Newborn Class. Parents who attend the class are given an additional packet of written information summarizing the topics covered in the Newborn Class.

The class begins by addressing the importance of brain development in the newborn period. Important concepts such as language development, parentese (talking to the baby using a singsong voice with words that are sophisticated and grammatically correct), sports casting (describing actions verbally to the baby) and labeling (commenting on the baby’s actions and reactions to what is happening around them) are explained and demonstrated by the pediatrician. The idea of responsive parenting is thoroughly described and demonstrated to the parents. The class then addresses common how-to medical questions: feeding frequency and amount; bowel movements and urination; burping and spitting up; umbilical cord care and bathing; diapering and circumcision care; skin and fingernail care; keeping baby from getting sick; colic and soothing the baby; and the importance of sleep and safe sleep habits. The pediatrician also addresses Baby Blues and Postpartum Depression, emphasizing the importance of support during the postpartum period. Finally, the pediatrician also reviews normal physical findings and exam findings from head to toe, and discusses when to call the doctor. The entire class is interactive and utilizes accessories, such as a doll, diapers, and a tub, to demonstrate concepts. Parents are encouraged to ask questions at any point in time.

For further details and outline of the intervention, as well as online materials, visit the Mount Sinai Parenting’s website at https://parenting.mountsinai.org/. The PDF of the Newborn Class is provided in **Appendix B**.

### 2.5. Outcome measures

The primary outcomes of this study were mean differences in maternal knowledge on newborn care, anxiety levels and confidence levels between the intervention and control groups. The survey took 10 minutes to administer. Anxiety level was measured first to minimize confounding effects of study participation on maternal anxiety. Parenting confidence was then measured, followed by knowledge. Basic demographic information such as the mother’s age, education level and race were collected at the end of the survey. Given the availability of many parenting interventions, we also asked if the mother had received other interventions outside of Mount Sinai. In the intervention group, mothers were also asked if they found the Newborn Class to be useful. Qualitative comments of what was and was not helpful were also elicited. The survey and answer key for the knowledge scale are provided in **Appendix A**.

#### 2.5.1. Maternal anxiety

Parental anxiety is a common measure of the well-being of mothers and a predictor of parenting behaviors. A widely used instrument for measuring anxiety levels is the State-Trait Anxiety Inventory for Adults (STAI-AD) (31)(32)(33). This self-administered scale was constructed by Spielberger et al (34) and has been translated into more than 30 languages for use in cross-cultural clinical studies. The scale differentiates between temporary “state anxiety” and more enduring “trait anxiety”. State anxiety is more responsive to change and thus a better measure of immediate post-intervention effect of the class. Given the overwhelming burdens the participants already have as new mothers, we aimed to limit the overall survey time. Therefore, only the state anxiety portion of the scale was used for the study, which consists of 20 brief statements describing how the participants feel at that moment in time. 10 items reflected positive states (e.g. I feel calm; I feel satisfied), and 10 items reflected negative states (e.g. I feel anxious; I feel indecisive). Four response options were given: (1) not at all; (2) somewhat; (3) moderately so; and (4) very much so. Mothers completed this portion of the survey independently. The possible range of scores is 10-40, with higher scores indicating higher levels of anxiety. License to reproduce the scale was purchased from the official distributor of STAI-AD, Mind Garden, Inc. and can be found in **Appendix C**.

#### 2.5.2. Maternal confidence

Maternal confidence was measured using the standardized Karitane Parenting Confidence Scale (KPCS), a scale developed for mothers or fathers of infants aged 0-12 months (35) and commonly used in parent-child intervention studies (36)(37)(38). The scale consists of 15 statements that are task-specific. Four response options were given: (1) hardly ever; (2) not very often; (3) some of the time; and (4) most of the time. The possible range of score is 0-45, with higher scores associated with higher levels of confidence. This scale was administered in the style of an interview, which is consistent with how the scale was developed and intended to be used. Participants were given a laminated card with the four response options, and each statement was read out loud by the research assistant. Since many of the statements refer to parent-child interactions that mothers who just gave birth would not have had (e.g., taking care of a sick child), the research assistant asked the participants to answer these statements according to how confident they would feel if they were asked to carry out these actions now. Permission to use the scale was obtained from the Karitane Education Department and the scale can be found in its original format in **Appendix D**.

#### 2.5.3. Maternal newborn care knowledge

Assessment of maternal knowledge on newborn care is commonly conducted using intervention-specific scales (39). Therefore, a newborn care knowledge scale specifically targeting topics taught in the Newborn Class was developed by a team of 3 pediatricians and 2 specialists at Mount Sinai. The test consists of 10 true or false questions, covering both medical and developmental topics consistently covered in the Newborn Class. The score ranges from 0-10, with higher scores indicative of higher levels of parenting knowledge. This scale was also administered through interview and an “answer key” was provided to the parents at the end of the survey. The knowledge scale and answer key can be found in **Appendix A**.

### 2.6. Statistical analysis

Distribution of participant characteristics within and between the control and intervention groups were analyzed using linear regression analysis and χ2 tests, respectively. Primary outcome measures were knowledge, anxiety and confidence, and are reported as means after adjusting for the influence of covariates. STAI-AD consists of both positive and negative state items, and each state was also analyzed separately. Mean differences between control and intervention group were compared using two-tailed T-tests. Effect sizes for each of the three primary outcomes were carried out using Hedge’s g to indicate the magnitude of difference between intervention and control group. Hedge’s g is a measure of standardized mean difference often used with pretest posttest designs (40). Interpretation of effect sizes from a Hedge’s g test was compared to that of Cohen’s d, a similar test for larger sample sizes. For Cohen’s d, effect sizes between 0.2 and 0.3 are considered small, medium if between 0.4 and 0.7, and large if over 0.8 (41). Correlations between the primary outcomes were analyzed using Spearman’s rank correlation coefficient test. STAI-AD and KPCS are both standardized scales with normalized data, and both have validated clinically-severe cut-offs. A secondary outcome of the study was therefore to quantify the difference in proportion of mothers scoring in the clinically-severe ranges between control and intervention groups. A two-tailed Z-score test for two population proportions was carried out to compare the proportions in the two groups. For the knowledge test, an additional two-tailed Z-score test was carried out to compare differences in scores between developmental questions versus medical questions across the two groups.

## Results

### 3.1. Participant flow

A total of 360 mothers were screened for eligibility. Eighty-nine met all the inclusion criteria, but 5 mothers declined to take part in the survey. Of the 84 enrolled mothers, 36 (42.9%) were in the intervention group and 48 (57.1%) were in the control group. The consort flow diagram of the study participants is in Figure 1.

**Fig. 1.**
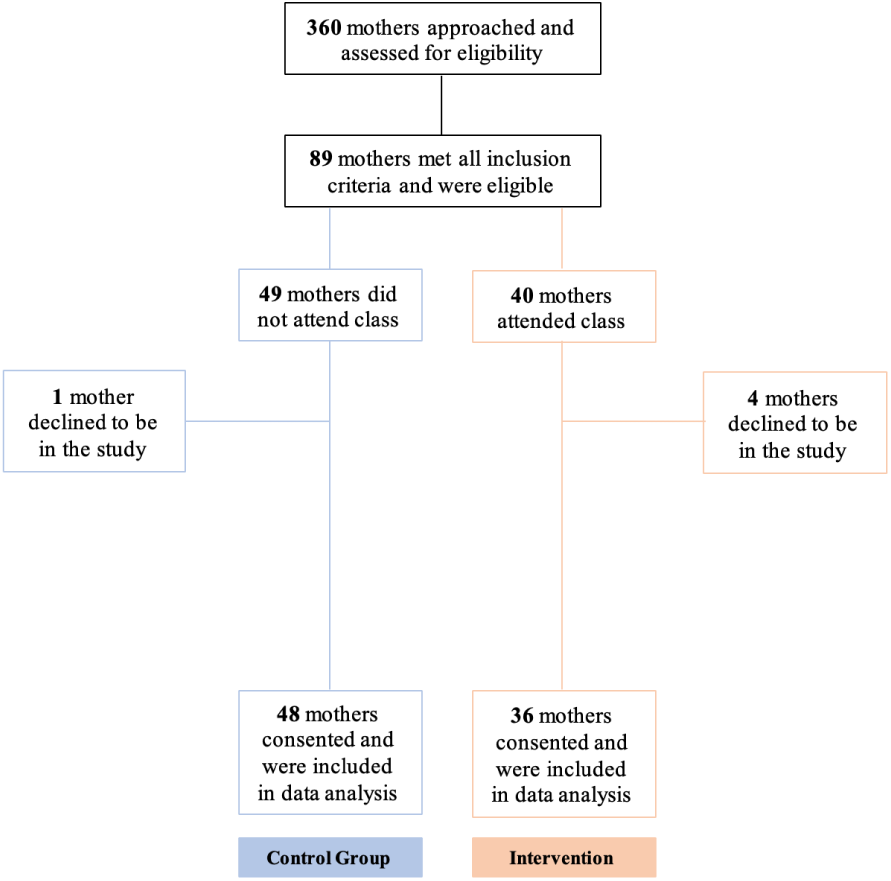
Consort Diagram. Flowchart showing the flow of participants through each stage of the trial. Three-hundred-and-sixty mothers were approached for the study. Out of the 89 mothers who met the inclusion criteria for the study, 5 declined to be in the study, 48 were included in the control group and 36 were included in the intervention group.

### 3.2. Demographic data

The mean (SD) age was 32.0 (4.34) years. Participants primarily self-identified as white (78.6%, n=66) and non-Hispanic (96.4%, n=81). Also, most participants were highly educated, reporting a master’s degree or higher (58.3%, n=49); only 6.0% reported high school diploma or below (n=5). However, chi-square analysis of the distribution of demographics showed no significant differences between control and intervention group (Table 1). Table 2 shows identified associations between demographic data and the three primary outcomes. As expected, there was a significant association between age and knowledge (Linear regression, *R*^2^ = 0.047, *P* = 0.049), with older individuals scoring better. However, age was not significantly associated with either confidence or anxiety. We also found a significant association between race and anxiety (Linear regression, *R*^2^ = 0.079, *P* = 0.010), with white participants exhibiting higher anxiety than non-white participants. Confidence was not associated with any demographic characteristic.

**Table 1.** Demographic characteristics. Basic demographic data collected from participants included age, highest level of education, race (white versus non-white) and ethnicity (Hispanic versus non-Hispanic). Chi-square analysis showed no significant differences were observed in any demographics between the intervention and control groups. Data are given as n (%).

| Categories | Control (n=48) | Intervention (n=36) | $\chi^2$ stat | p |
| --- | --- | --- | --- | --- |
|  | Value (%) | Value (%) |  |  |
| **Age ranges (years)** |  |  | 0.005 | 0.946 |
| 30 and below | 15 (31.3) | 11 (30.6) |  |  |
| 31 and above | 33 (68.7) | 25 (69.4) |  |  |
| **Education** |  |  | 1.5 | 0.472 |
| Highschool and below | 4 (8.3) | 1 (2.8) |  |  |
| Bachelors | 18 (37.5) | 12 (33.3) |  |  |
| Masters and above | 26 (54.2) | 23 (63.9) |  |  |
| **Race** |  |  | 0.849 | 0.357 |
| White | 36 (75) | 30 (83.3) |  |  |
| Non-White | 12 (25) | 6 (16.7) |  |  |
| **Ethnicity** |  |  | 0.72 | 0.396 |
| Hispanic | 1 (2.08) | 2 (5.56) |  |  |
| Non-Hispanic | 47 (97.92) | 34 (94.4) |  |  |

Significance level $\alpha = 0.05$

**Table 2.** Associations between primary outcomes. Linear regression analysis showed significant associations between parenting knowledge and maternal age, as well as between anxiety and race, with older participants showing higher knowledge and white participants exhibiting higher anxiety than non-white participants.

| Scale | Participants (n=84) $R^2$ [P-value] | | | |
| --- | --- | --- | --- | --- |
|  | Age | Education | Race | Ethnicity |
| **Knowledge** | **0.047 [0.049]** | 0.028 [0.126] | 0.043 [0.059] | 0.017 [0.238] |
| **Confidence** | 0.008 [0.406] | 0.035 [0.086] | 0.015 [0.266] | 0.004 [0.577] |
| **Anxiety** | 0.012 [0.328] | 0.035 [0.091] | **0.079 [0.010]** | 0.016 [0.901] |

Linear Regression Analysis.
Significance level $\alpha = 0.05$

### 3.3. Primary outcomes

See Table 3.

**Table 3.** Differences in knowledge, confidence and anxiety in control versus intervention mothers. Mothers in the intervention group exhibited significantly higher newborn care knowledge and confidence but no difference in overall anxiety. Interestingly, when the STAI-AD(S) items were separated into “positive” versus “negative” states, intervention mothers did show lower scores in positive state items, which translates to higher positive emotions such as “feeling calm”. Effect sizes were measured using Hedges’ g and compared to standardized Cohen d’s effect sizes used in similar studies; small effect: Hedge’s g between 0.2 and 0.3, medium effect: Hedge’s g between 0.4 and 0.7, large effect: Hedge’s g >0.8.

| Variables | Control (n=48) | Intervention (n=36) | Effect Size (Hedges' g) | p value |
| --- | --- | --- | --- | --- |
|  | Mean (SD) | Mean (SD) |  |  |
| Knowledge | 6.78 (1.25) | 8.08 (1.06) | 1.11 | <0.001 |
| Confidence | 35.20 (3.99) | 39.31 (3.88) | 1.03 | <0.001 |
| Anxiety | 38.50 (9.53) | 35.50 (8.73) | n/a | 0.164 |
| Positive State | 20.68 (5.28) | 18.00 (4.77) | 0.53 | 0.026 |
| Negative state | 17.80 (4.95) | 17.28 (4.64) | n/a | 0.642 |

SD: standard deviation; n/a: not assessed
Significance level $\alpha = 0.05$

#### 3.3.1. Knowledge

Mothers who attended the Newborn Class showed significantly higher levels of newborn care knowledge than participants in the control group (Two-tailed T-test, mean [SD], 8.08 [1.06] vs 6.78 [1.25]; P<0.001). A large effect size of 1.11 was observed when comparing the two groups.

#### 3.3.2. Confidence

A significantly higher level of confidence was observed for mothers in the intervention group compared to those in the control group (Two-tailed T-test, mean [SD], 39.31 [3.88] vs 35.20 [3.99]; P<0.001). A large effect size of 1.03 was observed.

#### 3.3.3. Anxiety

Although the intervention group showed lower maternal anxiety level, this difference was not statistically significant (Two-tailed T-test, mean [SD], 35.50 [8.73] vs 38.5 [9.53]; P=0.164).

### 3.4. Positive versus negative state-items on the anxiety scale

The STAI-AD(S) contains 20 brief statements describing how the participants feel at that moment in time. 10 items reflect positive states (e.g. I feel calm; I feel satisfied), and 10 items reflect negative states (e.g. I feel anxious; I feel indecisive). To evaluate if our intervention might preferentially target only one of these two types of states, we analyzed the scores of positive and negative state items separately. We found that although the intervention and control groups did not show an overall difference in maternal anxiety level, a statistically significant difference did emerge when the positive and negative state items were analyzed separately (Table 3). Specifically, the intervention group scored significantly lower on positive state items compared to the control group (Two-tailed T-test, mean [SD], 18.00 [4.77] vs 20.68 [5.28]; P=0.026). Because the STAI-AD(S) scores positive versus negative state items in opposing direction (higher score for reported negative items and lower score for reported positive items), this suggests that mothers in the intervention group felt more “calm” and “satisfied” without changes in “anxiety” and “indecisiveness”. A medium effect of 0.53 was observed for the positive state items. No significant difference was observed for the negative state items (Two-tailed T-test, mean [SD], 17.28 [4.64] vs 17.80 [4.95]; P=0.642).

### 3.5. Clinically severe ranges for anxiety and confidence levels

The STAI-AD (S) and KPCS can be used to detect clinically severe and moderate levels of anxiety and confidence derived from distributions of normalized data. A cutoff of 39-40 has been suggested to detect clinically significant symptoms for the STAI-AD (S) (42)(43). A moderate clinical range for KPCS is defined as scores between 31-35, and a severe clinical range as scores less than 31 (33). Two-tailed Z-score tests for two population proportions were used to compare the differences in proportion of mothers scoring in the clinically severe and moderate ranges between the intervention group and control group. About twice as many mothers in the control group scored in the clinically moderate KPCS range as compared to the intervention group (19.4% in intervention group versus 40.0% in control group, Two-tailed Z-score=-1.99, P=0.047). There was also a trend toward a decreased in incidence of mothers meeting clinically severe ranges for KPCS in the intervention group, though this decrease did not reach significance (2.8% in intervention group versus 15.5% in control group, Two-tailed Z-score=-1.84, P=0.066). The rates of clinically severe range scores in STAI-AD(S) did not differ significantly (30.6% in the intervention group versus 47.5% in control group, Two-tailed Z-score=-1.51, P=0.131). Outcomes from the two-tailed Z-score tests for anxiety and confidence are reported in Table 4.

**Table 4.** Proportions of participants scoring in clinical ranges for the KPCS and STAI-AD(S). A significantly lower proportion of mothers in the intervention group scored in the clinically moderate range of the KPCS. No group differences were observed in proportions of participants scoring in clinically severe ranges of the KPCS and STAI-AD(S).

| Cut-off Ranges | Control (n=48) | Intervention (n=36) | Z | p |
| --- | --- | --- | --- | --- |
| KPCS Clinically Moderate* | 18 (40.0%) | 7 (19.4%) | -1.99 | 0.047 |
| KPCS Clinically Severe* | 6 (15.5%) | 1 (2.8%) | -1.84 | 0.066 |
| STAI-AD (S) Clinically Severe** | 19 (47.5%) | 11 (30.6%) | -1.51 | 0.131 |

\*KPCS scores below 31 are considered in the severe clinical range; scores between 31-35 are in the moderate clinical range.
STAI-AD (S): State-Trait Anxiety Inventory - Adults (State items only); KPCS: Karlane Parenting Confidence Scale (blue=control group; red=intervention group).
\*\*STAI-AD scores above 39-40 has been suggested to detect clinically significant symptoms.
Significance level $\alpha = 0.05$

**Table 5.**
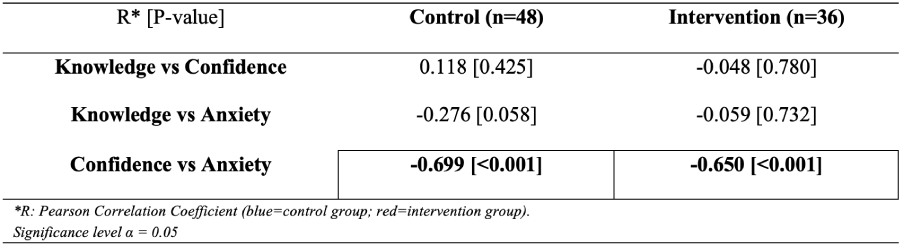
Correlations between primary outcomes. Control and intervention mothers showed similar profiles in correlations between primary outcomes. Knowledge did not correlate with either confidence or anxiety in either group. Consistent with prior studies, confidence was inversely related to anxiety and this relationship was similar in both intervention and control groups.

### 3.6. Correlations between primary outcomes

To assess potential associations among knowledge, anxiety and confidence, Pearson correlation coefficients were calculated within each group between primary outcomes. Significant negative correlations between anxiety and confidence were observed in both groups (Control group: Pearson correlation r=-0.699, p<0.001; Intervention group: Pearson correlation r=-0.650, p<0.001). However, knowledge was not correlated with anxiety or confidence in either group, and it is therefore an unlikely mediator of the higher confidence scores observed in the intervention group.

### 3.7. Differences in effect sizes on medical versus developmental newborn care topics

The Newborn Class was designed to cover both medical and developmental issues. The knowledge test created for this study thus includes questions that assess knowledge in both areas. To investigate if particular topics were preferentially retained by the mothers who attended the Newborn Class, differences in the proportion of correct answers was investigated for each question. Significantly higher proportion of mothers in the intervention group correctly answered three medical-type questions (Two-tailed Z-score test, Z=2.95, P=0.003; Z=2.95, P=0.016; Z=3.74, P<0.001) and one developmental-type question (Two-tailed Z-score test, Z=2.88, P=0.004) correctly. Results from the Z-score test are reported in Table 6.

**Table 6.**
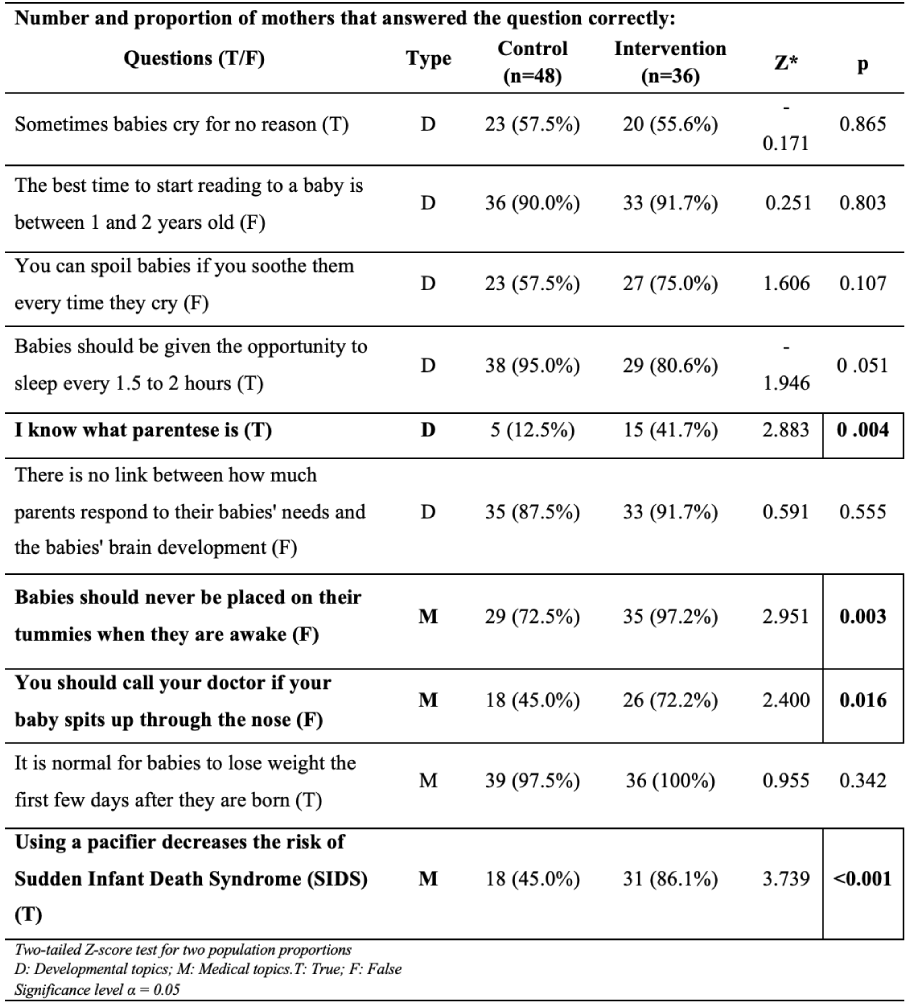
Knowledge scale. The knowledge test was specifically designed to assess concepts consistently covered in the Newborn Class. The test consisted of ten “true” (T) or “false” (F) type statements, six of which assessed developmental-type knowledge (D) and four of which assessed medical-type knowledge (M). Correct answers are shown in parenthesis following each statement. Proportion of mothers in control versus intervention groups correctly answering each knowledge question was compared using Two-tailed Z-score tests for two populations. The intervention significantly improved the knowledge of a total of four questions, one of which was developmental-type and three of which were medical-type.

## Discussion

In this nonrandomized controlled trial, we found that mothers who attended a pediatrician-led Newborn Class during their postpartum hospital stay scored significantly higher in knowledge and parenting confidence compared to mothers in the control group. The intervention was associated with a large effect size in both knowledge and confidence. Interestingly, while the overall anxiety score did not differ between the two groups, a significantly lower anxiety score was observed in the intervention group for the positive state items of STAI-AD (S). Lower score in the positive anxiety items translates to improved positive emotions (examples of positive emotion in the STAI-AD(S) are feeling “at ease”, “satisfied” and “relaxed”). Conceivably, improved knowledge about caring for their baby enhanced positive emotions of mothers in the intervention group without decreasing negative emotions (examples of negative emotions in the STAI-AD(S) are feeling “upset”, “tense” and “nervous”). Overall, our data indicate that a one hour-long pediatrician-led parenting intervention is effective in changing several outcomes associated with positive parenting.

It is important to consider that the control group had access to the same information as the intervention group on Mount Sinai’s online video database. While the study did not directly evaluate the number of mothers in the control group that had accessed these videos at the time of survey administration, our data clearly point to the importance of direct physician-mother interaction. During the Newborn Class, the teaching pediatrician conveys the information in an engaging manner through live demonstrations. The pediatrician also encourages parents to ask questions and engages them in conversation.

One of the inclusion criteria for the control group was answering positively to the intent to attend the class if it were available during the mother’s postpartum stay. Therefore, a potential selection bias could have arisen from the apprehensive-participant role, in which participants answer positively to please the researcher (43). It is unlikely however, that selection bias explains the entirety of the observed variance. Differences were observed in confidence and knowledge, which are directly targeted in the Newborn Class, but were not significant in levels of overall anxiety, which is not the focus of the class. The well-described inverse relationship between confidence and anxiety previously observed in the literature (44)(45)(46) was replicated here, and was of similar magnitude for both the intervention and the control groups. Additionally, the lack of correlation between knowledge – which measures topics specific to the Newborn Class – and either confidence or anxiety in either group, argues against a direct mediating effect of acquired knowledge on the higher confidence level in the intervention group. More likely, this improvement in confidence is mediated by direct physician-mother interaction. Finally, the overall similarity in demographics and similar profiles in the correlations between primary outcomes in control versus intervention groups, also argue against selection bias as a major contributor to the main effects observed in the intervention group. Nevertheless, an important future step for the verification of the effectiveness of the Newborn Class will be to conduct a randomized clinical trial, and/or pre-/post-intervention measures.

A surprising finding was the larger effect on medical over developmental knowledge. Developmental concerns are generally less addressed by pediatricians (47). Given the tremendous importance of promoting healthy brain development for establishing developmental trajectories, addressing these topics is of fundamental importance in pediatrics. With this in mind, the Newborn Class was designed to teach parents both medical and developmental topics regarding the care for their children. Therefore, we expected the unique emphasis on developmental issues to be reflected in selective increase in knowledge of these topics. However, we found the opposite: the class significantly enhanced the knowledge of three out of four medical topics but only one out of six developmental topics. A possible explanation for this is the larger knowledge deficit of developmental topics among new mothers. A well described phenomenon in human memory is that retention of new information is dependent on the number of times the information is encountered (48). Because medical concerns are more widely known, preexisting awareness of these topics might make it easier to consolidate the information during the Newborn Class. If so, it is imperative that we design methods to repeatedly remind parents of their important role in promoting their baby’s brain development.

## Limitations

This is an initial evaluation of the immediate effects of attending a novel Newborn Class developed by the Mount Sinai Parenting Center, and the results should be interpreted with prudence. Our findings demonstrate significant improvement in maternal parenting confidence and newborn care knowledge after participating in the class. Whether these immediate effects can be sustained will require longitudinal follow-up studies. Because of our strict inclusion/exclusion criteria, which were designed to increase validity in a study with relatively small sample size, it is currently unknown if the findings will generalize to e.g. multiparous mothers, mothers who experienced complications during the pregnancy, and mothers who delivered via cesarean section. Furthermore, this study only assesses the effect of the New-born Class on mothers. Fathers play a key role in parenting and significantly influence a child’s developmental trajectory (49)(50). Since the Newborn Class is taught to both parents, future studies evaluating the effect of the intervention on the fathers will be imperative. Additionally, our sample population was predominately white, educated, older mothers. This surprising and unintentional selection bias was likely a consequence of the strict inclusion/exclusion criteria and might further limit the generalizability of our findings. However, it should be noted that there was no significant difference in the demographics of the intervention and control groups. Therefore, at least in this population, our findings are likely meaningful. Most importantly, the ability of interventions, such as the Newborn Class, to actually alter developmental outcomes of babies will need to be evaluated in future studies.

## Conclusions

This study is the first to evaluate the Newborn Class at Mount Sinai Hospital. Results shows that this short pediatrician-led parenting intervention taught during the mothers’ postpartum stay in the hospital can be effective in significantly increasing maternal knowledge on newborn care and maternal confidence level. The Newborn Class can be easily implemented into other postpartum settings. Further research is needed to assess the long-term effects of the Newborn Class and the role of mothers’ confidence and knowledge on optimizing the developmental trajectories of their children.

## Data Availability

There are no linked research data sets, however raw data will be provided upon request.

## ACKNOWLEDGEMENTS

We are extremely grateful to all the mothers who participated in this study. Special thanks to the staff of The Lauder Center for Maternity Care at the Klingen-stein Pavilion of Mount Sinai Hospital for their help throughout the research.

